# The association between work situation and life satisfaction during the COVID-19 pandemic: prospective cohort study in Norway

**DOI:** 10.1101/2020.12.16.20248321

**Authors:** Ellen Øen Carlsen, Ida Henriette Caspersen, Helga Ask, Ragnhild Eek Brandlistuen, Lill Trogstad, Per Magnus

**Author notes:** Correspondence to: Ida Henriette Caspersen, Centre for Fertility and Health, Norwegian Institute of Public Health, P.O. Box 222 Skøyen, 0213 Oslo, Norway.

## Abstract

**Objectives:** To analyse the population effects on life quality of being laid off from work, having to work from home, or having been diagnosed with COVID-19.

**Design:** Nationwide population-based cohort study.

**Setting:** Norway.

**Participants:** We followed more than 80,000 participants in an ongoing cohort study, the Norwegian Mother, Father and Child Cohort Study (MoBa) during the COVID-19 pandemic. We analysed current life satisfaction in April and again in September/October 2020 for subjects whose work situation and infection status had changed.

**Main outcome measures:** Self-reported satisfaction with life, using a scale from 0 (worst) to 10 (best).

**Results:** Temporary and permanent layoffs, working from a home-based office, and getting a COVID-19 diagnosis were associated with modestly, but significantly lower concurrent life satisfaction, both on a population level and for subjects who changed status. The associations with change in work situation were stronger for men. For men with permanent job loss, the adjusted odds ratio for low life satisfaction (<6) was 3.2 (95% CI 2.4 to 4.2) in April and 4.9 (95% CI 3.5 to 6.9) in autumn. A suspected or confirmed COVID-19 diagnosis was associated with an adjusted odds ratio for low life satisfaction of 1.9 (95% CI 1.6 to 2.3) in spring. The strength of associations between work situation and life satisfaction did not vary much across socio-economic strata, but layoffs were more common among those with low education.

**Conclusion:** Layoffs, home office and infection status had clear impact on the quality of life as measured with a global life satisfaction scale. These findings suggest that social differentials in quality of life, are increasing during the pandemic.

**Funding:** This work was funded by the Norwegian Research Council’s Centres of Excellence Funding Scheme (no. 262700) and by the Norwegian Institute of Public Health (NIPH).

**SUMMARY BOXES:** *What is already known on this topic:*

− Being laid off from work or having to work from a home-based office is usually associated with reduced life quality.
− The population effect has not been estimated during the present surge in cases of COVID-19 in Europe.

*What this study adds:*

− This population-based study shows that life satisfaction in Norway has been stable from the first to the second wave of the pandemic, but that both layoffs and working from home is associated with reduced life satisfaction, especially among men.
− The reduced life satisfaction in people working from a home-based office implies that large proportions of the population are affected.

## INTRODUCTION

The COVID-19 pandemic has led to an increase in permanent and temporary layoffs.(1, 2) Studies from spring 2020 suggest that young people and those with low income and educational level were most often affected.(1, 3) Mental health and psychological well-being has decreased during the pandemic, in line with findings from previous pandemics or major lockdown situations, leading to concern about the global impact on mental health.(4-6) A German study found overall decreased satisfaction with work and family life during lockdown.(7) A study from Cyprus found increased levels of anxiety and depression among those who were unemployed, but no clear findings with regard to working from home compared to normal working days.(8) A study from China suggested that people who stopped working as a consequence of the pandemic reported worse mental and physical health conditions.(9) There is a need to estimate more precisely the effects of both being temporarily or permanently laid off from work and the effect of working from home. To arrive at population-based estimates, we examine these associations in a large, ongoing cohort, where participants were recruited several years prior to the pandemic. Our aim was to describe the level of self-rated life satisfaction during the spring and autumn of 2020, contrasting people with a stable work situation to people with moderate (home office) or major changes (permanent or temporary layoff in either spring or autumn). Similarly, we describe changes in life satisfaction following a COVID-diagnosis. We also aimed to assess whether associations varied across various socio-economic measures.

## METHODS

### Study design

The Norwegian Mother, Father and Child Cohort Study (MoBa)(10) is an ongoing nationwide cohort study in which 95,000 pregnant women and 75,000 partners were recruited between 1999 and 2008. Parents and children have been followed with questionnaires and registry linkages with the aim to understand causes of disease. The current sub-study was approved by The Regional Committee for Medical and Health Research Ethics, South East Norway C, no. 127708.

### Participants

Since March 2020, MoBa parents, now aged 30 to 65 years, have been invited to answer short mobile-phone questionnaires every 14 days regarding symptoms related to COVID-19, chronic illnesses, job situation, life satisfaction, and more. About 149,000 parents, who were still active MoBa participants, were invited to answer the repeated surveys, in this paper referred to as rounds. We used questionnaire rounds 2 and 3 (April 2020) and 14 and 15 (September/October 2020), including questions on both work situation and life satisfaction. Data from the various rounds was linked to previous MoBa data to obtain information on educational level and previously reported levels of life satisfaction. The participation rates were 74%, 68%, 58% and 58%, for rounds 2, 3, 14 and 15, respectively.

### Outcomes

Our main outcome was the participants’ global judgement of their own life, referred to as life satisfaction, measured on an adapted version of the Cantril ladder.(11) Participants were asked to rate their life at the moment on a scale from 0 to 10, with 0 representing the worst possible life and 10 being the best. We used life satisfaction both as a continuous variable (original and z-score) and as a binary variable, where a score of 6 or more was labelled ‘High Life Satisfaction’ and less than 6, ‘Low Life Satisfaction’.

### Exposures

The main exposure was participants’ response to the item ‘have you experienced change in work situation due to the COVID-19 pandemic’ categorized as ‘No’ or ‘Yes’. In autumn, the response alternatives were extended to include the categories ‘No’, ‘Yes-previously’ or ‘Yes-still change in work situation’. If ‘yes’ the answers were further grouped into loss of job, layoff from job, home-based office, or other change. A secondary exposure was self-reported COVID-19 diagnosis in spring (round 2 or 3), or in the period from late spring to autumn (rounds 4-14), based on participants answer to ‘have you tested positive for COVID-19’ or if they reported to have a suspected or confirmed COVID-19 diagnosis from their physician.

### Covariates

Age was categorized in 5- and 10-year intervals. We used information on educational level (less than high school, high school, university/college up to and including 4 years, more than 4 years of university/college). For men, this was reported in 2015 or at recruitment, and if not available, from the maternal recruitment questionnaire, where she reported on her partner’s educational attainment. For the women, educational level was extracted from a questionnaire sent out 8 years after recruitment or from the recruitment questionnaire. From previous MoBa data collections, we had information on their answers to the standardized Life Satisfaction Scale.(12) This scale includes the following 5 items: In most ways my life is close to my ideal; The conditions of my life are excellent; I am satisfied with my life; So far, I have had the important things I want in life; If I could live my life over, I would change almost nothing. Each item is scaled from 1 to 7, where 1 is “disagree completely” and 7 is “agree completely”. The mean value across the responses to all items was calculated, to obtain a score between 1 and 7. For women, the scale was responded to during pregnancy, as well as 3, 5 and 8 years after childbirth, while men answered in the recruitment questionnaire as well as in a questionnaire issued in 2015. We constructed an individual base-level life satisfaction score by calculating the mean across all available assessments of the Life Satisfaction Scale. As an alternative approach, we also calculated the base-level life satisfaction score using the most recent available assessment, which was adjusted for number of years since this was reported. From the COVID-19 questionnaires, we also obtained information on the number of persons in their household, and whether they suffered from any of a series of listed chronic disease (coded as dummy variables for each chronic disease).

### Statistical analysis

We estimated population-level associations between work situation or infection status and life satisfaction during spring and autumn using linear and logistic regression models. Multivariable regression analyses were adjusted for age-group, chronic diseases, number of persons in the household, educational level, as well as their mean score on the previously recorded Life Satisfaction Scale. As a sensitivity analysis, we adjusted for the most recent life satisfaction measure and number of years since this was reported, which did not change our findings (results not shown). We also performed analyses stratified by gender, educational level and age group. To investigate how a change in work situation or infection status was associated with life satisfaction on an individual level, we compared person-specific changes in life satisfaction z-scores from spring to autumn in groups who did or did not experience a change in work situation or infection status. The analyses were performed using R, version 4.0.2.

### Role of the funding source

The funders of the study had no role in study design, data collection, data analysis, data interpretation, or writing of the report. They did not participate in the decision to submit the manuscript for publication. All authors had full access to all data and were responsible for the decision to submit the manuscript for publication.

## RESULTS

Men’s life satisfaction was similar in spring and autumn 2020, while women appeared to have slightly higher life satisfaction in the autumn (Table 1). The overall proportion with low life satisfaction score decreased from 16% and 13% in the two survey rounds in spring to 12% in the autumn survey. The proportion of participants who reported to have lost their jobs was stable (<1%) during the study period, while the proportions who reported temporary layoff and use of home-based office substantially declined from spring to autumn: from 8% to 1% for layoff and from 35% to 13% for home-based office (Table 1). The proportion of participants reporting suspected/confirmed COVID-19 was 0.9% in spring (rounds 2-3). In the autumn rounds, another 0.9% of the respondents reported suspected/confirmed COVID-19 to have occurred (rounds 4-14).

**Table 1:** Descriptive characteristics of the study population.

|  | April 14,<br>2020 | April 29,<br>2020 | September 30,<br>2020 | October 14,<br>2020 |
| --- | --- | --- | --- | --- |
| **Characteristics** | **Round 2** | **Round 3** | **Round 14** | **Round 15** |
| **Participants, no.** | 109870 | 101744 | 85433 | 86337 |
| **Men, no. (%)** | 45344 (41) | 41281 (41) | 33238 (39) | 33876 (39) |
| **Women, no. (%)** | 64526 (59) | 60463 (59) | 52195 (61) | 52461 (61) |
| **Age (years), no. (%)** |  |  |  |  |
| 25-34 | 1110 (1) | 975 (1) | 637 (1) | 659 (1) |
| 35-39 | 9951 (9) | 8909 (9) | 6764 (8) | 6862 (8) |
| 40-44 | 30786 (28) | 28478 (28) | 23399 (27) | 23563 (27) |
| 45-49 | 40412 (37) | 37577 (37) | 32064 (38) | 32375 (37) |
| 50-54 | 21124 (19) | 19726 (19) | 17186 (20) | 17420 (20) |
| 55-59 | 5216 (5) | 4888 (5) | 4308 (5) | 4353 (5) |
| 60+ | 1271 (1) | 1191 (1) | 1075 (1) | 1105 (1) |
| **Any chronic disease, no. (%)<sup>1</sup>** | 30984 (28) | 28304 (28) | 24985 (29) | 25236 (29) |
| **Educational level, no. (%)** |  |  |  |  |
| < High school | 7314 (7) | 6447 (6) | 4843 (6) | 5004 (6) |
| High school | 33122 (30) | 30097 (30) | 24230 (28) | 24713 (29) |
| College ≤4 years | 38799 (35) | 36527 (36) | 31786 (37) | 31905 (37) |
| College >4 years | 25865 (24) | 24488 (24) | 21421 (25) | 21461 (25) |
| Missing/other | 4770 (4) | 4185 (4) | 3153 (4) | 3254 (4) |
| **Current work situation, no. (%)** |  |  |  |  |
| No/other change | 62252 (57) | 63255 (62) | NA | 73720 (85) |
| Home-based office | 38136 (35) | 30304 (30) | NA | 10981 (13) |
| Lay-off | 8653 (8) | 7477 (7) | NA | 1019 (1) |
| Loss of job | 637 (0.6) | 559 (0.5) | NA | 477 (0.6) |
| Missing | 192 (0.2) | 149 (0.1) | NA | 140 (0.2) |
| **Life satisfaction (0-10), mean (SD)** |  |  |  |  |
| Men | 7.3 (1.7) | 7.4 (1.6) | 7.4 (1.6) | NA |
| Women | 7.1 (1.7) | 7.3 (1.6) | 7.5 (1.6) | NA |
| **Life satisfaction, no. (%)** |  |  |  |  |
| Low (≤5) | 17458 (16) | 13035 (13) | 10184 (12) | NA |
| High (≥6) | 92150 (84) | 88521 (87) | 75139 (88) | NA |
| Missing | 262 (0.2) | 188 (0.2) | 110 (0.1) | NA |

<sup>1</sup> Information on chronic disease was collected in rounds 2 and 3 only. For rounds 14 and 15, proportions with chronic disease are calculated among those with available information from rounds 2 and 3 (10% had missing information about chronic disease both in round 14 and 15).

Temporary layoff or permanent job loss were more common among those with low education level than those with high educational level, both in the spring (Figure 1) and in the autumn (Supplementary Figure 1, Online Supplementary Material). Those with a high educational level (college or higher) reported most frequent use of home-based office (Figure 1). In autumn, overall proportions with home-based office were substantially reduced across gender, age groups and education level, but was now more common among men than women and among those with high education level.

**Figure 1.**
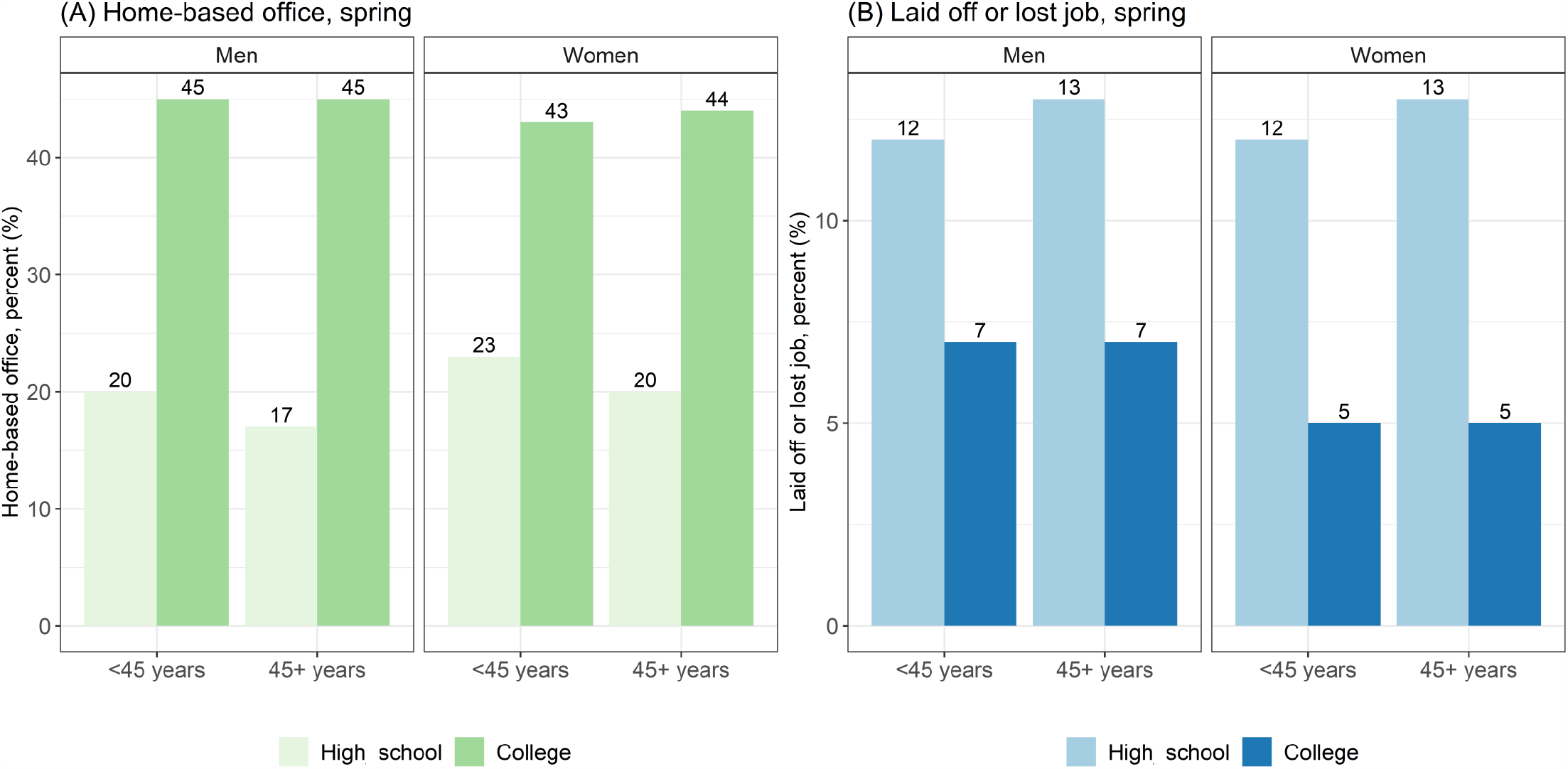
Proportions with (A) home-based office and (B) who were laid off (temporary) or lost job (permanently) in spring (survey round 2) across gender, age group and educational level.

Those who had permanently lost their jobs reported lower life satisfaction (Table 2), and their odds ratio of having low life satisfaction was increased in both genders when compared to those who had no or “other” change in their work situation (Table 3). For temporary layoffs, the results were similar, but the associations were of smaller magnitude than for those who had permanently lost their job. The magnitudes of associations between job loss or layoffs and reduction in life satisfaction were larger for men than women. Magnitudes of associations with life satisfaction were larger in autumn than in spring for layoffs in both genders, and for permanent job loss in women, but not in men.

**Table 2:** Regression analyses of mean life satisfaction (scale 0-10), stratified by survey round and gender.

| Exposure | Round 2 (spring) |  | Round 3 (spring) |  | Round 14/15 (autumn) |  |
| --- | --- | --- | --- | --- | --- | --- |
|  | Beta (95% CI),<br>unadjusted | Beta (95% CI),<br>adjusted* | Beta (95% CI),<br>unadjusted | Beta (95% CI),<br>adjusted* | Beta (95% CI),<br>unadjusted | Beta (95% CI),<br>adjusted* |
| **Men:** | n=42,861 |  | n=39,102 |  | n=25,274 |  |
| **Work situation** |  |  |  |  |  |  |
| No/other change (reference) | 0 (Ref) | 0 (Ref) | 0 (Ref) | 0 (Ref) | 0 (Ref) | 0 (Ref) |
| Home-based office | -0.2 (-0.3, -0.2) | -0.3 (-0.3, -0.3) | -0.2 (-0.3, -0.2) | -0.3 (-0.3, -0.2) | -0.3 (-0.3, -0.2) | -0.3 (-0.3, -0.2) |
| Lay-off | -0.5 (-0.6, -0.5) | -0.5 (-0.6, -0.5) | -0.6 (-0.6, -0.5) | -0.6 (-0.6, -0.5) | -1.0 (-1.2, -0.9) | -1.1 (-1.2, -0.9) |
| Loss of job | -1.0 (-1.2, -0.8) | -1.1 (-1.3, -0.9) | -1.5 (-1.7, -1.3) | -1.4 (-1.6, -1.2) | -1.6 (-1.8, -1.4) | -1.4 (-1.6, -1.2) |
| **Women:** | n=61,873 |  | n=58,222 |  | n=41,793 |  |
| **Work situation** |  |  |  |  |  |  |
| No/other change (reference) | 0 (Ref) | 0 (Ref) | 0 (Ref) | 0 (Ref) | 0 (Ref) | 0 (Ref) |
| Home-based office | -0.1 (-0.1, 0.0) | -0.1 (-0.1, -0.1) | -0.1 (-0.1, -0.1) | -0.2 (-0.2, -0.1) | -0.2 (-0.2, -0.1) | -0.2 (-0.3, -0.2) |
| Lay-off | -0.2 (-0.3, -0.2) | -0.2 (-0.3, -0.2) | -0.3 (-0.3, -0.2) | -0.3 (-0.3, -0.2) | -0.6 (-0.8, -0.5) | -0.7 (-0.8, -0.5) |
| Loss of job | -0.5 (-0.7, -0.4) | -0.6 (-0.8, -0.4) | -0.9 (-1.0, -0.7) | -0.9 (-1.0, -0.7) | -1.2 (-1.5, -1.0) | -1.1 (-1.3, -0.9) |

\*Adjusted for: age, educational level, chronic conditions, base-level life satisfaction, and number of people in household

**Table 3:** Odds ratios for having low life satisfaction (below 6) according to work situation, stratified by survey round and gender.

| Exposure | Round 2 (spring) |  | Round 3 (spring) |  | Round 14/15 (autumn) |  |
| --- | --- | --- | --- | --- | --- | --- |
|  | OR (95% CI),<br>unadjusted | OR (95% CI),<br>adjusted# | OR (95% CI),<br>unadjusted | OR (95% CI),<br>adjusted# | OR (95% CI),<br>unadjusted | OR (95% CI),<br>adjusted# |
| **<u>Men:</u>** | n=42,861 |  | n=39,102 |  | n=25,274 |  |
| **Work situation** |  |  |  |  |  |  |
| No/other change (reference) | 1 (Ref) | 1 (Ref) | 1 (Ref) | 1 (Ref) | 1 (Ref) | 1 (Ref) |
| Home-based office | 1.1 (1.0, 1.1) | 1.1 (1.1, 1.2) | 1.0 (0.9, 1.1) | 1.1 (1.0, 1.2) | 1.2 (1.1, 1.3) | 1.2 (1.1, 1.4) |
| Lay-off | 2.0 (1.8, 2.1) | 2.0 (1.8, 2.2) | 2.1 (1.9, 2.3) | 2.2 (2.0, 2.4) | 3.6 (2.8, 4.4) | 4.0 (3.1, 5.1) |
| Loss of job | 3.2 (2.5, 4.1) | 3.2 (2.4, 4.2) | 5.2 (4.0, 6.7) | 4.8 (3.6, 6.3) | 5.6 (4.1, 7.5) | 4.9 (3.5, 6.9) |
| **<u>Women:</u>** | n=61,873 |  | n=58,222 |  | n=41,793 |  |
| **Work situation** |  |  |  |  |  |  |
| No/other change (reference) | 1 (Ref) | 1 (Ref) | 1 (Ref) | 1 (Ref) | 1 (Ref) | 1 (Ref) |
| Home-based office | 0.9 (0.9, 1.0) | 1.0 (0.9, 1.0) | 1.0 (0.9, 1.1) | 1.0 (0.9, 1.0) | 1.1 (1.0, 1.1) | 1.1 (1.0, 1.2) |
| Lay-off | 1.4 (1.3, 1.5) | 1.4 (1.3, 1.5) | 2.1 (1.9, 2.3) | 1.5 (1.3, 1.6) | 2.2 (1.7, 2.7) | 2.4 (1.9, 3.0) |
| Loss of job | 2.1 (1.7, 2.6) | 2.2 (1.7, 2.8) | 5.2 (4.0, 6.7) | 2.8 (2.1, 3.6) | 3.8 (2.8, 5.1) | 3.7 (2.7, 5.0) |

For home-based office we found associations with reduced mean life satisfaction both in spring and autumn, but of smaller magnitude than for job loss and layoffs (Table 2). We found no clear associations between home-based office and low life satisfaction during spring. In the autumn, men with home-based office had an increased odds ratio of having low life satisfaction, and a similar but weaker trend was seen for the women (Table 3).

In analyses stratified by educational level, we found only small differences between low and high educational level in the odds ratios of low life satisfaction. For temporary layoff, odds ratios were slightly higher for those with college or higher education in spring, while for permanent job loss, odds ratios were slightly higher among those with high school or lower education (Supplementary Table 1, Online Supplementary Material). However, after restricting analyses to only those aged under 45 years, we found higher odds ratios for low life satisfaction among those with high education who had permanently lost their job, and the associations were even stronger in the autumn (Supplementary Table 2, Online Supplementary Material). When stratifying on both gender and age, the magnitudes of associations with low life satisfaction were highest in autumn among men above 45 years, followed by men in both age groups in spring, who had in both cases permanently lost their job (Supplementary Table 3, Online Supplementary Material).

Within individuals, difference in life satisfaction from spring to autumn were associated with changes in work situation (Figure 2). Those who were laid off in spring and were back to their normal working situation in autumn had an increased (0.13, 95% CI 0.10 to 0.16) life satisfaction z-score from spring to autumn. For those who were laid off in both spring and autumn, life satisfaction z-scores decreased (−0.12, 95% CI −0.19 to −0.05) from spring to autumn, while those with no change in work situation in either spring or autumn reported no change (−0.01 95% CI −0.02 to 0.0) in life satisfaction between spring and autumn.

**Figure 2.**
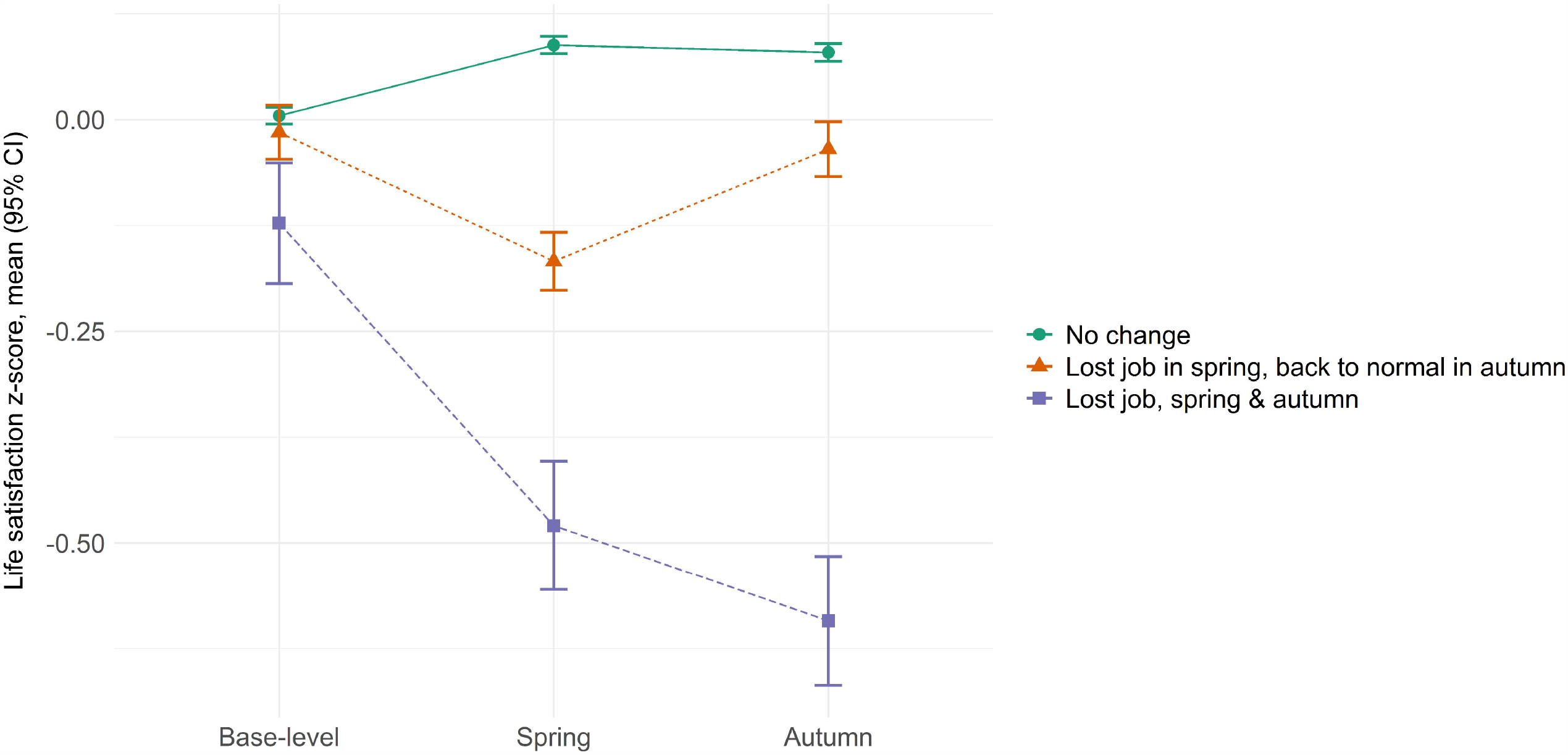
Satisfaction with life (mean z-score and 95% confidence intervals, CI) among those with no change in work situation in spring or autumn (n=39,280), those who had temporarily or permanently lost their job in spring (April, round 2 or 3), but were back to their normal work situation in autumn (September/October, n=3976); and those who reported to have lost their job in both spring and autumn (n=984).

In spring, those with a suspected (by physician) or confirmed (by testing) COVID-19 diagnosis had increased odds ratios (1.9, 95% CI 1.6 to 2.3) of low life satisfaction compared to those who did not report COVID-19 at all. In autumn, those who got a COVID-19 diagnosis in the period May-October had also increased odds ratio (1.7, 95% CI 1.3 to 2.0), although with weaker magnitude than those in spring, of low life satisfaction compared to those with no COVID-19 diagnosis in either spring or autumn (Table 4).

**Table 4:** Logistic and linear regression analyses of infection status (suspected or confirmed COVID-19) and life satisfaction stratified by survey round. Adjusted analyses adjusted for: chronic conditions, base-level life satisfaction, and number of people they cohabit with.

|  | Spring, round 2-3 |  | Autumn, round 4-14 |  | Spring, round 2-3 |  | Autumn, round 4-14 |  |
| --- | --- | --- | --- | --- | --- | --- | --- | --- |
|  | OR<br>(95% CI),<br>unadjusted | OR<br>(95% CI),<br>adjusted | OR<br>(95% CI),<br>unadjusted | OR<br>(95% CI),<br>adjusted | Beta<br>(95% CI),<br>unadjusted | Beta<br>(95% CI),<br>adjusted | Beta<br>(95% CI),<br>unadjusted | Beta<br>(95% CI),<br>adjusted |
|  | n=89,622 |  | n=73,416 |  | n=89,622 |  | n=73,416 |  |
| COVID-19 |  |  |  |  |  |  |  |  |
| No (reference) | 1 (Ref.) | 1 (Ref.) | 1 (Ref.) | 1 (Ref.) | 0 (Ref.) | 0 (Ref.) | 0 (Ref.) | 0 (Ref.) |
| Yes | 2.1 (1.8, 2.5) | 1.9 (1.6, 2.3) | 1.7 (1.4, 2.1) | 1.6 (1.3, 2.0) | -0.3 (-0.4, -0.3) | -0.5 (-0.6, -0.4) | -0.4 (-0.5, -0.3) | -0.3 (-0.5, -0.2) |

Changes in life satisfaction from spring to autumn was also related to infection status (Figure 3). Those who had reported a suspected or confirmed COVID-19 diagnosis in the period between spring and autumn, had a −0.22 (95% CI −0.32 to −0.11) reduction in life satisfaction z-score from spring to autumn. For those reporting COVID-19 in spring, life satisfaction z-score tended to increase from spring to autumn (0.10, 95% CI 0.01 to 0.20), while we found no change (0.0, 95% CI −0.01 to 0.0) in life satisfaction among those not reporting a COVID-19 diagnosis.

**Figure 3.**
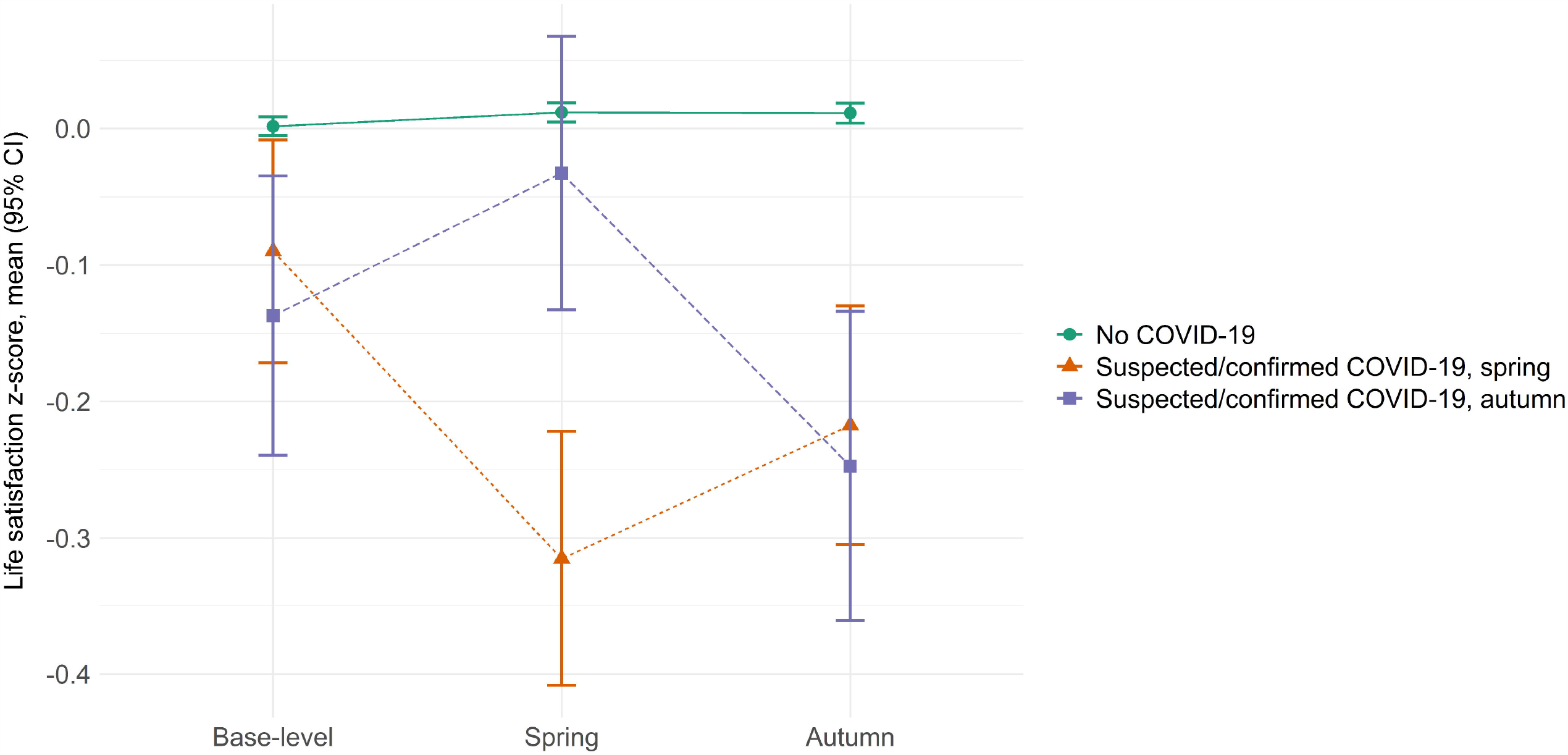
Satisfaction with life (mean z-score and 95% confidence intervals, CI) among those with no suspected/confirmed COVID-19 diagnosis (n=73,837), those who had suspected or confirmed COVID-19 in spring (round 2 or 3, n=844), and among those who reported in autumn that they had a suspected or confirmed COVID-19 diagnosis in the period between spring and autumn (new cases in round 4-14, n=422).

## DISCUSSION

In this large population-based study, we estimated life satisfaction levels as a consequence of the changing work situations and infection status during the first six months of the COVID-19 pandemic in Norway. We found clear indications of lower life satisfaction among those who experienced either a permanent or temporary job loss and among those who reported to be working from a home-based office. The associations were somewhat stronger for men. We found that the association between layoffs and reduced life satisfaction was stronger after the summer, when a lower proportion of participants reported to be laid off. Our findings indicate that associations with reduced life satisfaction were of larger magnitude for job loss than for getting a COVID-19 diagnosis.

A main strength of our study is the large sample size and continued high participation rates over all survey rounds. Both genders and participants from all parts of the country are well represented in the study, and there is a wide age span. Furthermore, the advantages of using pre-existing cohorts as opposed to panels recruited during the pandemic has been outlined as advantageous.(13) MoBa is a cohort that was recruited among pregnant women and their partners during 1999-2008. Thus, we do not know whether the association between job loss and life satisfaction is of the same magnitude in childless adults, although it seems unlikely that it would be very different. A study of the recruitment into MoBa indicated a higher socio-economic status among participants than the general Norwegian population.(14) However, the effect of job loss on life satisfaction in this study was about the same across levels of educational attainment.

Norway is a relatively generous welfare state, and it is likely that the gradients we find in reduced life satisfaction would have been stronger in countries where temporary and permanent layoffs are associated with less generous compensations. According to the OECD Economic Survey from 2016, Norwegians were in overall more satisfied with their lives than average in other OECD countries.(15) Life satisfaction for Norwegians in 2016 was also slightly higher (score 7.6) than for participants in the current study (7.1 to 7.5), which may indicate decreased overall life satisfaction in the population due to the pandemic.

Other studies have shown conflicting results regarding overall changes in population mental health and psychological well-being during the COVID-19 pandemic, but certain groups, such as younger age groups, may be more vulnerable.(4, 16, 17) Individuals rate their life satisfaction relative to their conception of what maximum and minimum satisfaction would be. In addition to a rather stable life satisfaction level rooted in personality, it is hypothesized that life satisfaction is achieved when various needs are met, and when engaging in meaningful activities.(18) A job can both fulfil needs of income, belonging and provide meaningful activities for individuals.

There is little available evidence on effects of mandatory home-based office on mental health. However, in a small study from China, life satisfaction decreased among those who stopped working due to the COVID-19 pandemic, but no differences were seen between those working from home versus from the office.(9) In a study from Germany, work satisfaction decreased among women who were being sent on short-time work, but no associations were found between home-based office and work or family satisfaction.(7)

Studies conducted before the pandemic have found that layoffs are associated with decreased life satisfaction.(19) In addition to affecting quality of life, unemployment has been suggested as a risk factor for morbidity and mortality, in particular for men,(20, 21) with an increased risk of myocardial infarction the first year after losing their job.(22) Whether the same applies in a situation where layoffs are more widespread due to a collective crisis such as the ongoing pandemic, remains to be elucidated. However, unemployment has been associated with increased risk of suicide even in the context of a collective financial crisis.(23) Although the incidence of ST-elevation myocardial infarctions dropped after the COVID-19 pandemic hit Northern Europe,(24) long-term health consequences might be substantial. Continued research into effects of changes in working situations on somatic and mental health are required to better inform healthcare personnel on how to follow-up on patients who have been affected by layoffs during the COVID-19 pandemic.

Our findings indicate that changes in work situation ranging from home-based office to permanent job loss during the COVID-19 pandemic impacts life satisfaction. Temporary layoff or permanent job loss were more common among those with low education level than those with high educational level and may thus widen social differentials in health. There is a need to follow population-based cohort studies through and after the pandemic for better estimates of long-term health consequences.

### KEY MESSAGES

Temporary and permanent layoffs, working from a home-based office, and getting a COVID-19 diagnosis were associated with lower concurrent life satisfaction, both on a population level and for subjects who changed status during April to October 2020.

## Data Availability

The data underlying this article are available for analysis after approval from a Norwegian ethics committee and application to the Norwegian Institute of Public Health.

## ACKNOWLEDGEMENTS

The Norwegian Mother, Father and Child Cohort Study is supported by the Norwegian Ministry of Health and Care Services and the Ministry of Education and Research. We are grateful to all the participating families in Norway who take part in this on-going cohort study.

## CONTRIBUTOR AND GUARANTOR INFORMATION

LT, PM designed the study and collected the data. EØC and IHC performed statistical analyses. EØC, REB, HA and IHC interpreted the data. EØC, IHC and PM wrote the first draft of the manuscript. LT, REB and HA revised the manuscript. IHC accept full responsibility of the work and controlled the decision to publish. All authors read and approved the final version.

## DECLARATION OF INTERESTS

All authors have completed the ICMJE uniform disclosure form at www.icmje.org/coi_disclosure.pdf and declare: no support from any organisation for the submitted work; no financial relationships with any organisations that might have an interest in the submitted work in the previous three years; no other relationships or activities that could appear to have influenced the submitted work.

## TRANSPARENCY STATEMENT

The authors affirm that this manuscript is an honest, accurate, and transparent account of the study being reported; that no important aspects of the study have been omitted; and that any discrepancies from the study as planned (and, if relevant, registered) have been explained.

## DATA SHARING

Data from the cohort is available for analysis after approval from a Norwegian ethics committee and application to the Norwegian Institute of Public Health.

## Notes

### Competing Interest Statement

The authors have declared no competing interest.

### Funding Statement

This work was funded by the Norwegian Research Council Centres of Excellence Funding Scheme (no. 262700) and by the Norwegian Institute of Public Health (NIPH).

### Author Declarations

The current sub-study was approved by The Regional Committee for Medical and Health Research Ethics, South East Norway C, no. 127708.

