## Supplementary material for "The association between work situation and life satisfaction during the COVID-19 pandemic: prospective cohort study in Norway": Online Supplementary Material

Supplementary Table 1. Logistic regression analyses of low life satisfaction stratified by survey round and education level. Adjusted analyses adjusted for: age, chronic conditions, base-level life satisfaction, and number of people they cohabit with.

| | Round 2 (spring) | | Round 3 (spring) | | Round 14/15 (autumn) | |
|---|---|---|---|---|---|---|
| | **OR low life satisfaction (95% CI), unadjusted** | **OR low life satisfaction (95% CI), adjusted** | **OR low life satisfaction (95% CI), unadjusted** | **OR low life satisfaction (95% CI), adjusted** | **OR low life satisfaction (95% CI), unadjusted** | **OR low life satisfaction (95% CI), adjusted** |
| **≤ High school** | n=39,566 | | n=35,833 | | n=22,558 | |
| **Work situation** | | | | | | |
| No/other change (reference) | 1 (Ref.) | 1 (Ref). | 1 (Ref.) | 1 (Ref.) | 1 (Ref.) | 1 (Ref.) |
| Home-based office | 1.0 (0.9, 1.1) | 1 (1.0, 1.1) | 0.9 (0.8, 1.0) | 0.9 (0.9, 1.0) | 1.1 (0.9, 1.3) | 1.1 (1, 1.3) |
| Lay-off | 1.4 (1.3, 1.6) | 1.5 (1.4, 1.6) | 1.5 (1.4, 1.7) | 1.6 (1.5, 1.8) | 2.7 (2.1, 3.3) | 3.0 (2.4, 3.7) |
| Loss of job | 2.8 (2.2, 3.5) | 2.9 (2.2, 3.7) | 3.3 (2.6, 4.3) | 3.4 (2.6, 4.5) | 4.5 (3.3, 6.2) | 4.3 (3.1, 5.9) |
| **College** | n=63,736 | | n=60,212 | | n=53,207 | |
| **Work situation** | | | | | | |
| No/other change (reference) | 1 (Ref.) | 1 (Ref.) | 1 (Ref.) | 1 (Ref.) | 1 (Ref.) | 1 (Ref.) |
| Home-based office | 1.0 (1.0, 1.1) | 1.0 (1.0, 1.1) | 1.0 (1.0, 1.1) | 1.0 (1.0, 1.1) | 1.2 (1.1, 1.3) | 1.2 (1.1, 1.3) |
| Lay-off | 1.8 (1.6, 1.9) | 1.8 (1.6, 1.9) | 1.8 (1.7, 2.0) | 1.9 (1.7, 2) | 2.7 (2.0, 3.4) | 2.9 (2.2, 3.8) |
| Loss of job | 2.4 (1.8, 3.1) | 2.3 (1.8, 3) | 3.6 (2.7, 4.7) | 3.4 (2.6, 4.5) | 4.4 (3.2, 5.9) | 4.1 (3.0, 5.6) |

Supplementary Table 2. Logistic regression analyses of low life satisfaction(score <6) in a subset with age<45 and educational level ≤high school stratified by survey round. Adjusted analyses adjusted for: chronic conditions, base-level life satisfaction, and number of people they cohabit with.

| | **Spring, round 2** | | **Spring, round 3** | | **Autumn, round 14/15** | |
|---|---|---|---|---|---|---|
| | **OR (95% CI), unadjusted** | **OR (95% CI), adjusted** | **OR (95% CI), unadjusted** | **OR (95% CI), adjusted** | **OR (95% CI), unadjusted** | **OR (95% CI), adjusted** |
| **Age <45 , <= High school** | n=17,301 | | n=15,410 | | n=8779 | |
| **Work situation** | | | | | | |
| No/other change (reference) | 1 (Ref.) | 1 (Ref.) | 1 (Ref.) | 1 (Ref.) | 1 (Ref.) | 1 (Ref.) |
| Home-based office | 0.9 (0.8, 1.0) | 1.0 (0.9, 1.1) | 0.8 (0.7, 0.9) | 0.8 (0.7, 1.0) | 1.0 (0.8, 1.3) | 1.0 (0.8, 1.3) |
| Lay-off | 1.3 (1.1, 1.4) | 1.3 (1.2, 1.5) | 1.4 (1.2, 1.6) | 1.5 (1.3, 1.7) | 2.1 (1.4, 3.1) | 2.1 (1.4, 3.1) |
| Loss of job | 3.4 (2.3, 5.0) | 3.6 (2.4, 5.3) | 3.2 (2.1, 4.8) | 3.3 (2.2, 5.0) | 3.3 (1.8, 5.9) | 2.9 (1.6, 5.3) |
| **Age <45 , College** | n=22,364 | | n=21,126 | | n=14,600 | |
| **Work situation** | | | | | | |
| No/other change (reference) | 1 (Ref.) | 1 (Ref.) | 1 (Ref.) | 1 (Ref.) | 1 (Ref.) | 1 (Ref.) |
| Home-based office | 1.0 (0.9, 1.1) | 1.0 (0.9, 1.1) | 1.0 (0.9, 1.1) | 1.0 (0.9, 1.1) | 1.1 (0.9, 1.3) | 1.1 (0.9, 1.3) |
| Lay-off | 1.7 (1.5, 2.0) | 1.7 (1.5, 2.0) | 1.5 (1.2, 1.8) | 1.5 (1.2, 1.8) | 1.4 (0.7, 2.5) | 1.3 (0.6, 2.4) |
| Loss of job | 2.6 (1.6, 4.2) | 2.4 (1.5, 3.9) | 4.6 (2.7, 7.4) | 4.2 (2.5, 7.0) | 5.5 (3.0, 9.7) | 5.3 (2.9, 9.6) |

Supplementary Table 3. Logistic regression analyses of low life satisfaction (score <6) stratified by survey round, gender, and age group. Adjusted analyses adjusted for: chronic conditions, base-level life satisfaction, and number of people they cohabit with.

| | **Spring, round 2** | | **Spring, round 3** | | **Autumn, round 14/15** | |
|---|---|---|---|---|---|---|
| | **OR (95% CI), unadjusted** | **OR (95% CI), adjusted** | **OR (95% CI), unadjusted** | **OR (95% CI), adjusted** | **OR (95% CI), unadjusted** | **OR (95% CI), adjusted** |
| **Men, <45 years** | n=13,079 | | n=11,701 | | n=6827 | |
| **Work situation** | | | | | | |
| No/other change (reference) | 1 (Ref.) | 1 (Ref.) | 1 (Ref.) | 1 (Ref.) | 1 (Ref.) | 1 (Ref.) |
| Home-based office | 1.0 (0.9, 1.1) | 1.0 (0.9, 1.2) | 1.1 (0.9, 1.2) | 1.1 (1.0, 1.3) | 1.1 (0.9, 1.3) | 1.1 (0.9, 1.3) |
| Lay-off | 2.1 (1.8, 2.4) | 2.0 (1.7, 2.4) | 2.2 (1.8, 2.6) | 2.1 (1.8, 2.5) | 3.2 (1.9, 5.2) | 2.6 (1.5, 4.2) |
| Loss of job | 5.0 (3.1, 8.1) | 5.2 (3.2, 8.5) | 5.3 (3.2, 8.5) | 5.0 (3.0, 8.3) | 4.4 (2.0, 9.0) | 3.7 (1.6, 7.8) |
| **Men, 45+ years** | n=29,782 | | n=27,401 | | n=18,447 | |
| **Work situation** | | | | | | |
| No/other change (reference) | 1 (Ref.) | 1 (Ref.) | 1 (Ref.) | 1 (Ref.) | 1 (Ref.) | 1 (Ref.) |
| Home-based office | 1.1 (1, 1.2) | 1.2 (1.1, 1.3) | 1.0 (0.9, 1.1) | 1.1 (1.0, 1.2) | 1.2 (1.1, 1.4) | 1.3 (1.2, 1.5) |
| Lay-off | 2.0 (1.8, 2.2) | 2.0 (1.8, 2.2) | 2.1 (1.9, 2.4) | 2.2 (1.9, 2.4) | 4.0 (3.1, 5.3) | 4.5 (3.4, 6.0) |
| Loss of job | 2.8 (2.0, 3.8) | 2.5 (1.8, 3.5) | 5.0 (3.6, 6.9) | 4.6 (3.3, 6.4) | 5.9 (4.1, 8.4) | 5.3 (3.6, 7.7) |
| **Women, <45 years** | n=27,007 | | n=25,194 | | n=16,745 | |
| **Work situation** | | | | | | |
| No/other change (reference) | 1 (Ref.) | 1 (Ref.) | 1 (Ref.) | 1 (Ref.) | 1 (Ref.) | 1 (Ref.) |
| Home-based office | 0.9 (0.8, 0.9) | 0.9 (0.9, 1.0) | 0.8 (0.8, 0.9) | 0.9 (0.8, 1.0) | 1.0 (0.8, 1.1) | 1.0 (0.9, 1.2) |
| Lay-off | 1.2 (1.1, 1.4) | 1.2 (1.1, 1.4) | 1.2 (1.1, 1.4) | 1.3 (1.1, 1.5) | 1.5 (1.0, 2.3) | 1.5 (0.9, 2.2) |
| Loss of job | 2.4 (1.6, 3.5) | 2.2 (1.5, 3.3) | 3.3 (2.2, 5.0) | 3.2 (2.1, 4.8) | 4.4 (2.7, 7.2) | 4.0 (2.4, 6.6) |
| **Women, 45+ years** | n=34,866 | | n=33,028 | | n=25,048 | |
| **Work situation** | | | | | | |
| No/other change (reference) | 1 (Ref.) | 1 (Ref.) | 1 (Ref.) | 1 (Ref.) | 1 (Ref.) | 1 (Ref.) |
| Home-based office | 0.9 (0.8, 0.9) | 0.9 (0.9, 1.0) | 1.0 (0.9, 1.1) | 1.0 (1.0, 1.1) | 1.1 (1.0, 1.2) | 1.2 (1.0, 1.3) |
| Lay-off | 1.2 (1.1, 1.4) | 1.2 (1.1, 1.4) | 1.6 (1.4, 1.8) | 1.7 (1.5, 1.9) | 2.8 (2.1, 3.6) | 3.0 (2.3, 4.0) |
| Loss of job | 2.4 (1.6, 3.5) | 2.2 (1.5, 3.3) | 2.6 (1.8, 3.5) | 2.6 (1.8, 3.6) | 3.8 (2.6, 5.5) | 3.5 (2.4, 5.1) |

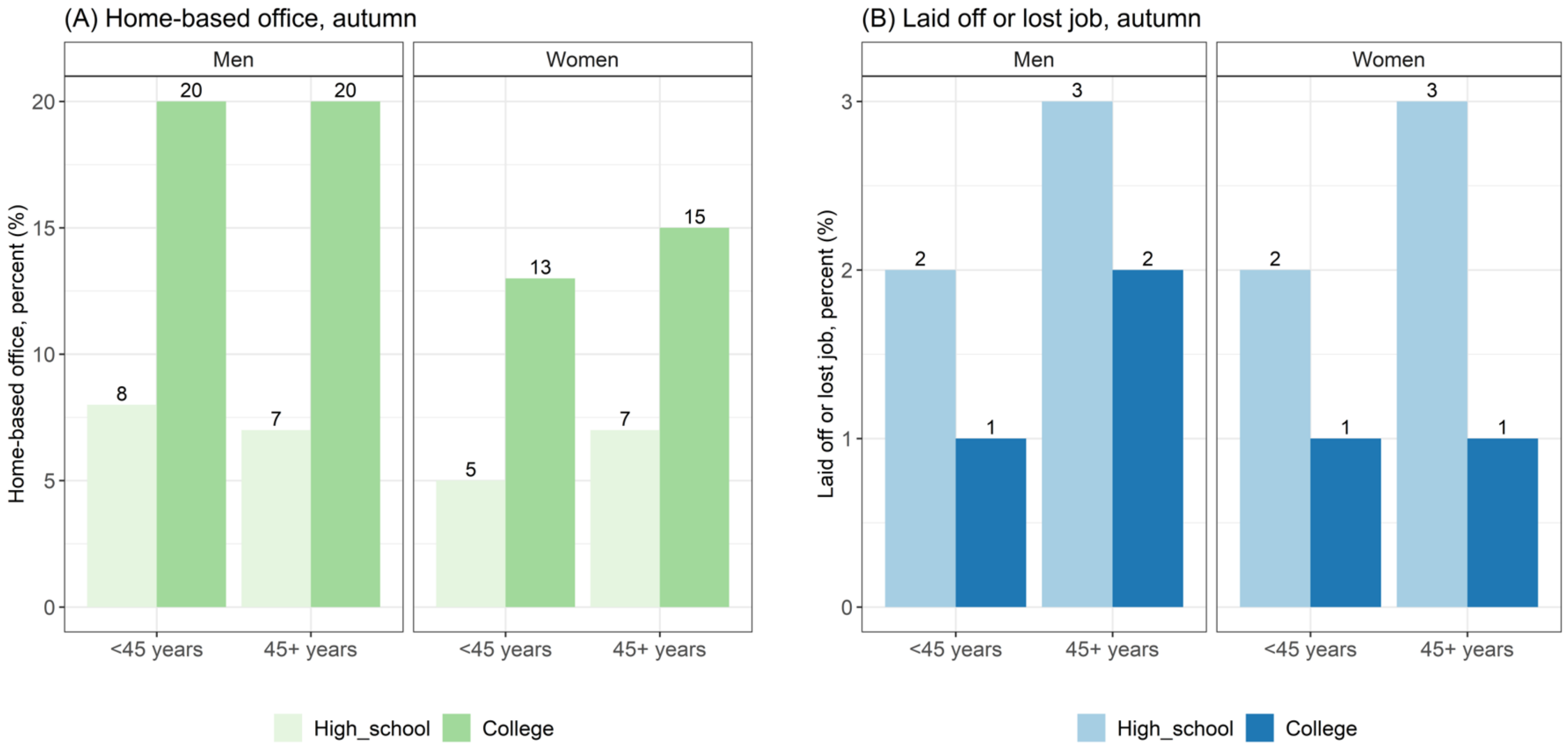


Supplementary Figure 1. Proportions with (A) home-based office and (B) who were laid off (temporary) or lost job (permanently) in autumn (survey round 15) across gender, age group and education level.
